# Connectome reorganization associated with temporal lobe pathology and its surgical resection

**DOI:** 10.1101/2023.11.13.23298482

**Authors:** Sara Larivière, Bo-yong Park, Jessica Royer, Jordan DeKraker, Alexander Ngo, Ella Sahlas, Judy Chen, Raúl Rodríguez-Cruces, Yifei Weng, Birgit Frauscher, Ruoting Liu, Zhengge Wang, Golia Shafiei, Bratislav Mišić, Andrea Bernasconi, Neda Bernasconi, Michael D. Fox, Zhiqiang Zhang, Boris C. Bernhardt

**Affiliations:** Multimodal Imaging and Connectome Analysis Laboratory, McConnell Brain Imaging Centre, Montreal Neurological Institute and Hospital, McGill University, Montreal, QC, Canada; Center for Brain Circuit Therapeutics, Brigham and Women’s Hospital, Harvard University, Boston, MA, USA; Department of Data Science, Inha University, Incheon, Republic of Korea; Center for Neuroscience Imaging Research, Institute for Basic Science, Suwon, Republic of Korea; Department of Medical Imaging, Jinling Hospital, Nanjing University School of Medicine, Nanjing, China; Analytical Neurophysiology Laboratory, Montreal Neurological Institute, McGill University, Montreal, QC, Canada; Department of Radiology, Nanjing Drum Tower Hospital, Affiliated Hospital of Nanjing University Medical School, Nanjing, China; Department of Psychiatry, Perelman School of Medicine, University of Pennsylvania, Philadelphia, PA, USA; McConnell Brain Imaging Centre, Montreal Neurological Institute and Hospital, McGill University, Montreal, QC, Canada; Neuroimaging of Epilepsy Laboratory, McConnell Brain Imaging Centre, Montreal Neurological Institute, McGill University, Montreal, QC, Canada

**Keywords:** Connectome, brain networks, focal lesion, surgery, longitudinal, gradients

## Abstract

Network neuroscience offers a unique framework to understand the organizational principles of the human brain. Despite recent progress, our understanding of how the brain is modulated by focal lesions remains incomplete. Resection of the temporal lobe is the most effective treatment to control seizures in pharmaco-resistant temporal lobe epilepsy (TLE), making this syndrome a powerful model to study lesional effects on network organization in young and middle-aged adults. Here, we assessed the downstream consequences of a focal lesion and its surgical resection on the brain’s structural connectome, and explored how this reorganization relates to clinical variables at the individual patient level. We included adults with pharmaco-resistant TLE (*n* = 37) who underwent anterior temporal lobectomy between two imaging time points, as well as age-and sex-matched healthy controls who underwent comparable imaging (*n* = 31). Core to our analysis was the projection of high-dimensional structural connectome data—derived from diffusion MRI tractography from each subject—into lower-dimensional gradients. We then compared connectome gradients in patients relative to controls before surgery, tracked surgically-induced connectome reconfiguration from pre-to postoperative time points, and examined associations to patient-specific clinical and imaging phenotypes. Before surgery, TLE presented with marked connectome changes in bilateral temporo-parietal regions, reflecting an increased segregation of the ipsilateral anterior temporal lobe from the rest of the brain. Surgery-induced connectome reorganization was localized to this temporo-parietal subnetwork, but primarily involved postoperative integration of contralateral regions with the rest of the brain. Using a partial least-squares analysis, we uncovered a latent clinical-imaging signature underlying this pre-to postoperative connectome reorganization, showing that patients who displayed postoperative integration in bilateral fronto-occipital cortices also had greater preoperative ipsilateral hippocampal atrophy, lower seizure frequency, and secondarily generalized seizures. Our results bridge the effects of focal brain lesions and their surgical resections with large-scale network reorganization and inter-individual clinical variability, thus offering new avenues to examine the fundamental malleability of the human brain.

## Introduction

Human brain organization is increasingly conceptualized and analyzed from a network perspective, allowing great strides in understanding both health and disease. In recent years, these approaches have been guided by advances in neuroimaging acquisition and complex data analytics. In particular, the advent of diffusion MRI tractography has enabled the approximation of structural connections *in vivo*, and the systematic characterization of brain connectivity—so called *connectomes*.^1^ In parallel, a series of multivariate analytics has been developed to capture organizational principles of large-scale brain connectivity. These range from graph theoretical assessments of network topology^2–4^ to modular decompositions that identify interacting systems.^5–7^ More recently, dimensionality reduction methods that derive compact and continuous embeddings from high dimensional connectomes have begun to reveal salient spatial axes of connectivity.^8,9^ In the current work, we adopted the latter approach to establish how structural connectivity embeddings can be modulated by focal lesions.

We specifically study patients with pharmaco-resistant temporal lobe epilepsy (TLE), who suffer from seizures originating from pathological alterations in the temporal lobe despite anti-seizure medical treatment.^10^ Compared to patients with adequate seizure control, pharmaco-resistant patients present with elevated risk for comorbidity and mortality,^11^ impaired quality of life and wellbeing,^12^ cognitive dysfunction and psychiatric difficulties,^13–16^ as well as progressive brain damage.^17–19^ In these patients, randomized controlled trials have shown that resection of the affected temporal lobe is currently the most effective treatment to control seizures, to restore psychosocial functioning, and to improve quality of life.^20,21^ By applying dimensional decomposition approaches to structural connectivity data acquired in healthy individuals as well as patients before and after surgery, we can investigate the impact of focal lesions on connectome-level organization.

Our novel framework, thereby, complements previous investigations of structural connectivity alterations in TLE patients relative to controls before surgery, which have reported microstructural and architectural damage seldom limited to the temporal lobe or to the hemisphere ipsilateral to the seizure focus.^22–25^ Similarly, prior work has assessed topological changes based on graph theoretical parameterization, and has accumulated evidence for a modular rearrangement in networks that extends beyond the temporal lobe.^26–30^ Such findings are compatible with the notion that “focal” pathologies generally affect large-scale brain connectivity, and that large-scale compromise contributes to patient presentation and clinically relevant outcomes.^31–35^ In line with these findings, studies assessing brain network changes following epilepsy surgery suggest that a focal resection does not only impact the primary lesioned site, but rather has effects that cascade along direct and indirect connections into mutually interconnected networks.^36–38^ As such, TLE represents a powerful human lesion model to assess how focal pathology and its resection affect whole-brain organization.

Here, we leveraged pre-as well as postoperative diffusion MRI data acquired in a cohort of pharmaco-resistant TLE patients. Our analytical paradigm derived whole brain connectomes from diffusion tractographic modelling, followed by non-linear dimensionality reduction to derive a continuous coordinate system in which confluent spatial trends of structural connectivity variation can be investigated.^8,39^ We compared our patients to a group of healthy controls, to investigate preoperative alterations and we tracked connectome reorganization from pre-to postoperative time points in patients. Exploiting inter-patient heterogeneity, we furthermore examined associations of pre-to postoperative connectivity trends with clinical and other imaging phenotypes at the individual patient level.

## Methods

### Participants

We studied 37 consecutive people suffering from pharmaco-resistant TLE, who (*i*) underwent anterior temporal lobectomy as a treatment of their seizures at Jinling Hospital between 2009 and 2018, (*ii*) had postoperative histological confirmation of hippocampal sclerosis, (*iii*) underwent a research-dedicated, high-resolution 3T MRI before and after surgery that included T1-weighted (T1w) and diffusion MRI scans, and (*iv*) had at least 1 year of postoperative follow-up. Patients were diagnosed according to the classification of the International League Against Epilepsy based on a comprehensive examination that includes clinical history, seizure semiology, continuous video-electroencephalographic (EEG) telemetry recordings, neuroimaging, and neuropsychology. No patient had encephalitis, malformations of cortical development (*e.g.*, tumors, vascular malformations), or a history of traumatic brain injury. We also studied 31 age-and sex-matched healthy individuals who underwent identical 3T MRI at one time point. Demographic and clinical information on both cohorts is provided in **Table S1**. Our study was approved by the research ethics board of Jinling Hospital, Nanjing University School of Medicine, and written informed consent was obtained from all participants.

### MRI acquisition and processing

All MRI data were obtained on a Siemens Trio 3T scanner (Siemens, Erlangen, Germany) and included: (*i*) high-resolution 3D T1w MRI using a magnetization-prepared rapid gradient-echo sequence (repetition time [TR] = 2300 ms, echo time [TE] = 2.98 ms, flip angle = 9°, voxel size = 0.5 × 0.5 × 1 mm^3^, field of view [FOV] = 256 × 256 mm^2^, 176 slices) and (*ii*) diffusion MRI using a spin echo-based echo planar imaging sequence with 4 b0 images (dMRI, TR = 6100 ms, TE = 93 ms, flip angle = 90°, FOV = 240 × 240 mm^2^, voxel size = 0.94 × 0.94 × 3 mm^3^, b-value = 1000 s/mm^2^, diffusion directions = 120).

Multimodal MRI preprocessing was performed using micapipe, an open-access image processing and data fusion pipeline (*v*0.1.4; https://micapipe.readthedocs.io).^40^ In brief, native T1w structural images were deobliqued, reoriented to standard neuroscience orientation, corrected for intensity nonuniformity, intensity normalized, skull-stripped, and submitted to FreeSurfer (*v*6.0)^41^ to extract models of the cortical surfaces. Subcortical structures were segmented using FSL FIRST.^42^ Native DWI images were denoised, underwent b0 intensity normalization, and were corrected for susceptibility distortion, head motion, and eddy currents. Left and right hemispheric data of individuals with TLE were sorted into ipsilateral/contralateral to the seizure focus. To minimize confounds related to normal inter-hemispheric asymmetries, prior to sorting we *z*-scored data in patients relative to controls.

### Surgical cavity mapping

Patient-specific surgical cavities were automatically segmented by registering both pre-and postoperative T1w images to the MNI152 standard template through linear transformations and subtracting the postoperative scan from the preoperative scan. Segmented cavities were visually inspected and manually edited to ensure that the extent of the resections was correctly identified.

Cavities were then mapped to the surface template, and a consensus label was generated as defined by the union of all segmentations.

### Structural connectome generation

Structural connectomes were generated with MRtrix3 from pre-processed DWI data.^43^ We performed anatomically-constrained tractography using tissue types (cortical and subcortical grey matter, white matter, cerebrospinal fluid) segmented from each participant’s pre-processed T1w images registered to native DWI space. We estimated multi-tissue response functions, performed constrained spherical-deconvolution and intensity normalization, generated a tractogram with 40M streamlines (maximum tract length = 250; fractional anisotropy cut-off = 0.06), and applied spherical deconvolution informed filtering of tractograms (SIFT2) to reconstruct whole-brain streamlines weighted by cross-sectional multipliers. Reconstructed streamlines were mapped onto 400 similar-sized cortical parcels constrained within the boundaries of the Desikan-Killiany atlas^44,45^ and 14 regions in the subcortex and the hippocampus to produce subject-specific structural connectivity matrices. Group-average normative structural connectomes were defined using a distance-dependent thresholding procedure, which preserved the edge length distribution in individual patients,^46^ and were log transformed to reduce connectivity strength variance. As such, structural connectivity was defined by the number of streamlines between two regions (*i.e.*, fiber density). Structural connectomes were thresholded to retain only the top 25% of connections.

### Connectome gradient estimation

Cortex-and subcortex-wide structural connectome gradients were generated using BrainSpace (*v*0.1.10; https://brainspace.readthedocs.io).^45^ First, a group-level structural connectome (combining cortical and subcortical data) was obtained using an iterative, leave-one-out procedure (**Figure 1**). For both the group-level and the left-out subject-specific connectome, an affinity matrix was constructed with a normalized angle kernel, and eigenvectors were estimated via diffusion map embedding, an unsupervised nonlinear dimensionality reduction technique that projects connectome features into low-dimensional gradients.^9,47^ This technique is only controlled by only few parameters, computationally efficient, relatively robust to noise compared to other nonlinear techniques,^48^ and has been extensively used in the previous literature.^8^ Algorithm parameters were identical to those from prior applications, specifically α = 0.05 and diffusion time *t* = 0 to retain the global relations between data points in the embedded space and to emphasize direct connections.^9^ Interhemispheric connections were not included in the gradient computation; left and right gradients were generated separately and aligned with Procrustes.^45^ The eigenvectors estimated from this technique provide a connectivity coordinate system—a diffusion map—where Euclidean distances in the manifold correspond to diffusion times between the nodes of the network.^47^ In this gradient space, regions with similar connectivity profiles are closely located, whereas regions with different connectivity profiles are located farther apart. To analyze gradient changes within and across participants, each left-out subject-specific gradient was aligned to the group template gradient (estimated using data from all other participants) via Procrustes alignment,^45^ normalized, and sorted into ipsilateral/contralateral to the seizure focus. The scree plot describing the eigenvalue decay function revealed that four principal eigenvectors (*i.e.*, gradients) corresponded to the cut-off point, after which eigenvalues leveled off at small values and only explained marginal information.

**Figure 1.**
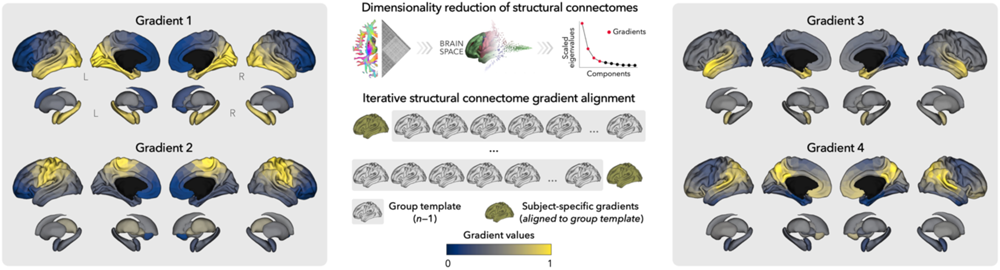
Construction of structural connectome gradients. Subject-specific structural connectome gradients were generated from diffusion MRI data and iteratively aligned to a group template (using data from all other participants). The first four gradients were retained.

### Quantifying pre-and postoperative changes in structural gradients

We assessed cross-sectional connectome changes between individuals with TLE (preoperatively) and controls using univariate fixed effects models performed on each eigenvector independently.^49^ Eigenvector-specific score histograms were then generated across the entire cortex, and split into ipsilateral/contralateral as well as patients/controls to examine changes in the distribution of significant scores and to assess inter-hemispheric asymmetry. To signify an overall load of alterations, we also compared the aggregate of the first four eigenvectors in patients relative to controls using a multivariate fixed effect linear model. By statistically combining multidimensional gradients, the latter approach leverages their covariance to obtain a substantial gain in sensitivity, and thus unveil subthreshold properties not readily identified in a single eigenvector. Both uni-and multivariate linear models tested for differences between groups while controlling for effects of age and sex using BrainStat (*v*0.4.2; https://brainstat.readthedocs.io).^49^

To test for changes over time (*i.e.*, pre-*vs*. postsurgery), we fitted univariate and multivariate linear mixed effects models, a flexible framework for the analysis of repeated measures.^49,50^ Our models controlled for effects of age and sex, and included a subject-specific random intercept to improve model fit in longitudinal designs. With this approach, we were able to test for pre-to postoperative within-subject individual and multivariate eigenvector changes.

Findings from all fixed and mixed effects models were corrected for multiple comparisons using the false discovery rate (FDR) procedure.^51^

## 4D gradient deformations

To simplify multidimensional changes into a single deformation scalar feature, we computed the Euclidean distance between the group template center and each subject-specific data point in the 4D gradient space (Figure 2B). We assessed patterns of eigenvector contraction (*i.e.*, connectome integration) and expansion (*i.e.*, connectome differentiation) relative to the centre of the gradient space. The template center was defined as the centroid of the first four eigenvectors. Shifts in structural connectivity patterns lead to a displacement of eigenvector scores in the 4D gradient space, which in turn affects their proximity to the center. Gradient contraction/expansion thus quantifies global brain reorganization in connectivity space.^52,53^ Gradient deformation was quantified only in regions of interest, as determined by areas that showed significant cross-sectional and longitudinal changes, respectively. The rationale was to determine whether preoperative and surgery-induced 4D gradient deformations were observed in areas that showed significant eigenvector changes.

**Figure 2.**
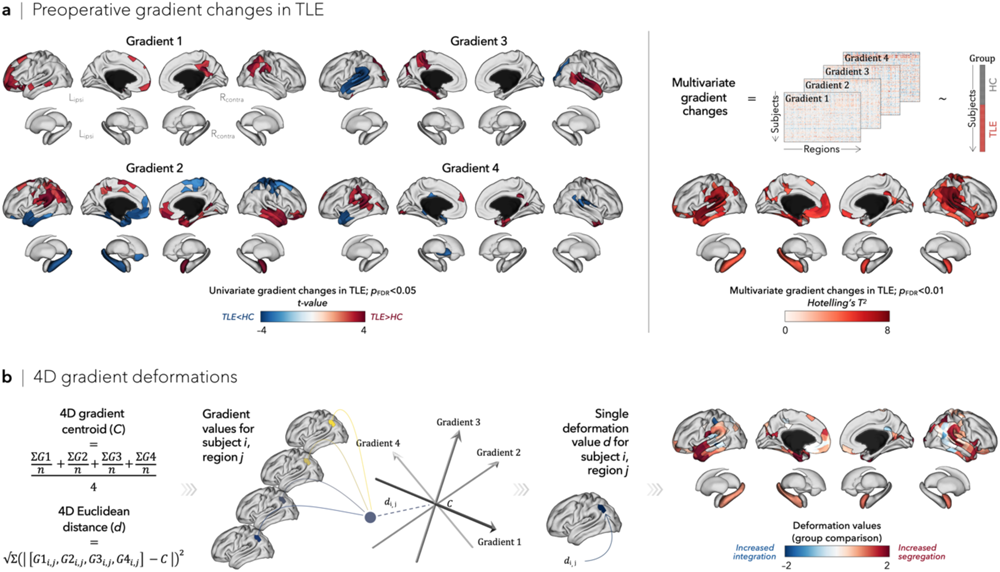
Preoperative structural connectome gradient changes in TLE. (**a**) Univariate (left) and multivariate (right) fixed effect models compared gradient values in individuals with TLE (before surgery) and healthy controls. Significant gradient changes in patients relative to controls were observed in bilateral orbitofrontal and temporoparietal cortices, ipsilateral hippocampus, and contralateral amygdala (all *p*_FDR_ < 0.05). (**b**) Multivariate gradient changes were simplified into a scalar feature of connectome deformations by computing the Euclidean distance (*d*_i,j_) between the group template center (*C*) and each subject-specific data point in the 4D gradient space. Restricting this analysis to areas of significant gradient changes identified in (**a**) revealed increased segregation of temporo-parietal regions in TLE relative to controls.

### Spatial associations between preoperative and surgery-induced reorganization

To assess whether preoperative alterations constrained surgery-induced changes, we performed spatial associations between cross-sectional (relative to controls) and longitudinal (pre-to postoperative) connectome findings. Statistical significance of spatial correlations was assessed using variogram-matching models that take into account the spatial dependencies in the data.^45,54,55^ This method generates surrogate brain maps with matched spatial autocorrelation to that of a target brain, and has previously been applied to cortical and subcortical structures. As recommended,^54^ surrogate maps were generated using surface-based geodesic distance between cortical regions and three-dimensional Euclidean distance between subcortical and cortical/subcortical regions. Variogram-matching null distributions were then generated from randomly shuffling surrogate maps while preserving the distance-dependent correlation between elements of the brain map. The empirical correlation coefficients were compared against the null distribution determined by the ensemble of spatially permuted correlation coefficients to estimate a *p*-value (termed *p*_var_).

### Clinical associations

We used partial least squares (PLS) analysis to unravel within-patient relationships between surgery-induced 4D gradient deformations and standard neuroimaging (*i.e.*, hippocampal atrophy^56^) and clinical parameters (Figure 4A). PLS analysis is a multivariate associative technique that maximizes the covariance between two sets of variables.^57^ Briefly, these two variable sets are correlated with each other across patients. The resulting correlation matrix is submitted to singular value decomposition, which identifies linear combinations of the original variables to generate new latent variables that have maximum covariance.^58^ We evaluated the significance of our model using nonparametric methods: (*i*) permutation tests were used to assess statistical significance of each latent variable (10,000 permutations; termed *p*_perm_), (*ii*) bootstrap ratios were used to assess the reliability of singular vector weights (analogous to *z*-scores, such that 95% confidence interval corresponds to a bootstrap ratio of ±1.96 and ±2.58, respectively), and (*iii*) cross-validation was used to assess out-of-sample correlations between projected scores (using 100 randomized train-test splits of the original data, where 75% of the data was treated as a training set and 25% of the data was treated as an out-of-sample test set). Mathematical details of the analysis and inferential methods are described elsewhere.^57,59^

## Results

### Connectome gradient analysis at baseline

For every participant, we generated cortex-and subcortex-wide structural connectome gradients using non-linear dimensionality reduction.^45^ The first four gradients explained approximately 75.28 ± 1.89% (baseline/preoperatively) and 78.85 ± 1.92% (postoperatively) of variance across participants, with each gradient representing a different axis of connectivity variation (Figure 1). Across both time points and in both groups, gradients generally depicted smooth differentiation in structural connectivity profiles of frontal *vs* temporo-occipital regions (*Gradient 1*), orbitofrontal *vs* sensorimotor regions (*Gradient 2*), medial occipital *vs* anterior temporal lobe regions (*Gradient 3*), and sensory *vs* transmodal regions (*Gradient 4*).

Compared to healthy controls, presurgical TLE patients showed significant gradient changes in bilateral temporo-parietal (*p*_FDR_ < 0.001) and orbitofrontal (*p*_FDR_ < 0.005) cortices, together with the ipsilateral hippocampus (*p*_FDR_ < 0.01) and the contralateral amygdala (*p*_FDR_ < 0.005; Figure 2A). Considering each gradient separately, global histogram analysis revealed significant asymmetry between ipsilateral and contralateral gradient alterations in TLE. Most notably, ipsilateral mesiotemporal lobe regions were shifted toward the most extreme, segregated positions along Gradients 2, 3, and 4, whereas their contralateral homologues were shifted to a more central, integrated position (**Figure S1**). This ipsilateral mesiotemporal lobe segregation was further confirmed by quantifying their positional change in the 4D gradient space relative to controls. Constraining this analysis to areas of significant preoperative gradient changes, individuals with TLE showed highest deformation values (*i.e.*, depicting increased expansion/segregation) in ipsilateral anterior temporal lobe (*p*uncorr < 0.005) and parahippocampal gyrus (*p*_uncorr_ < 0.01; Figure 2B).

### Tracking surgery-induced deformations

Directly comparing structural connectome changes before and after anterior temporal lobectomy (Figure 3A), patients showed significant multivariate gradient changes in posterior temporal regions near the resection site (*p*_FDR_ < 0.05) and contralateral temporo-parietal cortices (*p*_FDR_ < 0.05; Figure 3B). As evidenced in both univariate and 4D gradient deformation analyses, contralateral temporo-parietal regions were shifted towards more integrated positions in the 4D gradient space after surgery, particularly along the axis of Gradient 1 (*p*_FDR_ < 0.05; **Figure S2**).

**Figure 3.**
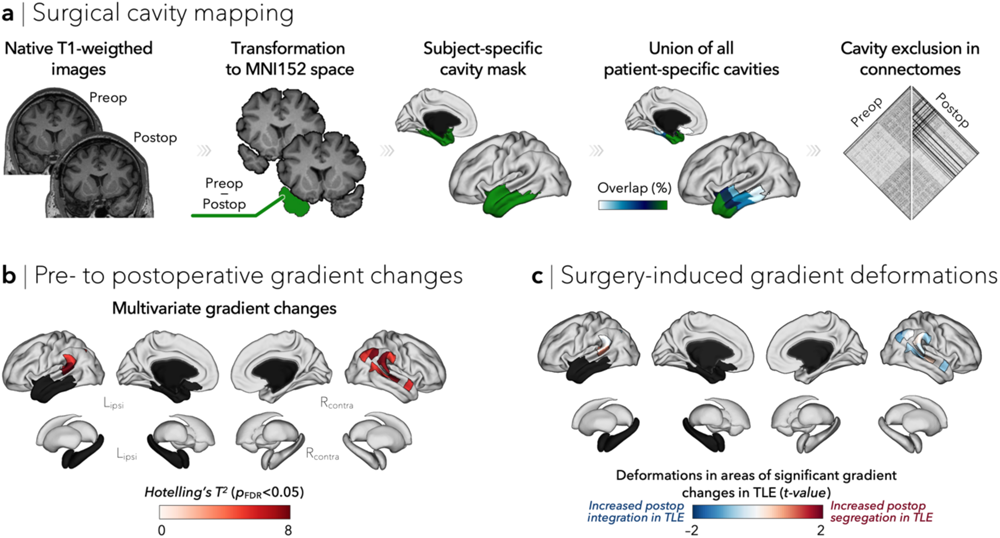
Surgery-induced structural connectome gradient changes in TLE. (**a**) Surgical cavities were automatically segmented from pre-and postoperative T1w MRIs. Cavity masks were excluded *a posteriori* in preoperative analysis and *a priori* in postoperative analyses. (**b**) Multivariate mixed effects models revealed a pattern of surgery-induced gradient changes encompassing ipsilateral posterior temporal and contralateral temporo-parietal regions (all *p*_FDR_ < 0.05). (**c**) Analysis of 4D gradient deformations constrained to areas of multivariate gradient change revealed segregation of ipsilateral temporal regions but increased integration of contralateral temporo-parietal regions after surgery.

As all regions showing pre-to postoperative gradient changes fell within the boundaries of significant preoperative changes, we next assessed the extent to which preoperative connectome reorganization relates to surgery-induced changes. The degree of preoperative multivariate gradient changes was correlated with pre-to postoperative multivariate gradient changes across all regions (*r* = 0.74, *p*_var_ < 0.0001), as well as when considering only areas of significant preoperative (*r* = 0.72, *p*_var_ < 0.0001) and pre-to postoperative (*r* = 0.49, *p*_var_ = 0.05) gradient changes (**Figure S3**). In other words, regions with profound connectome alterations before surgery (relative to controls) corresponded to regions that also underwent the largest pre-to postoperative reorganization. We also found a significant, albeit weaker, negative spatial associations between whole-brain preoperative and surgery-induced 4D gradient deformations (*r* = –0.15, *p*_var_ < 0.005). In this case, a negative correlation indicates that regions that were initially segregated from the rest of the brain before surgery subsequently shifted towards a more integrated position in the connectome space after surgery.

### Associations with clinical variables

Multivariate PLS analysis identified one statistically significant latent variable (LV-1 accounting for 44.65% of covariance, *p*_perm_ < 0.05) that maximized the covariance between whole-brain 4D surgery-induced deformation patterns and standard neuroimaging parameters (*e.g.*, hippocampal atrophy) and clinical measures (Figure 4A). Loadings (*i.e.*, correlations of individual clinical measures with LV-1) revealed ipsilateral hippocampal atrophy in CA1-3 and dentate gyrus subfields (all *r* < –0.28), lower seizure frequency (*r* < –0.22), and presence of secondarily generalized seizures (*r* > 0.35) as the strongest contributors of LV-1. Patients presenting with these clinical features showed increased postoperative connectome integration in bilateral fronto-occipital cortices, ipsilateral postcentral gyrus, and contralateral mesiotemporal cortex (all bootstrap ratio < 95% confidence interval). Clinical measures and 4D surgery-induced gradient deformations were correlated at the individual patient level (*r* = 0.52), while out-of-sample correlations averaged *r* = 0.20 (and exceeded mean permuted out-of-sample correlations of *r* = – 0.028; Figure 4B).

**Figure 4.**
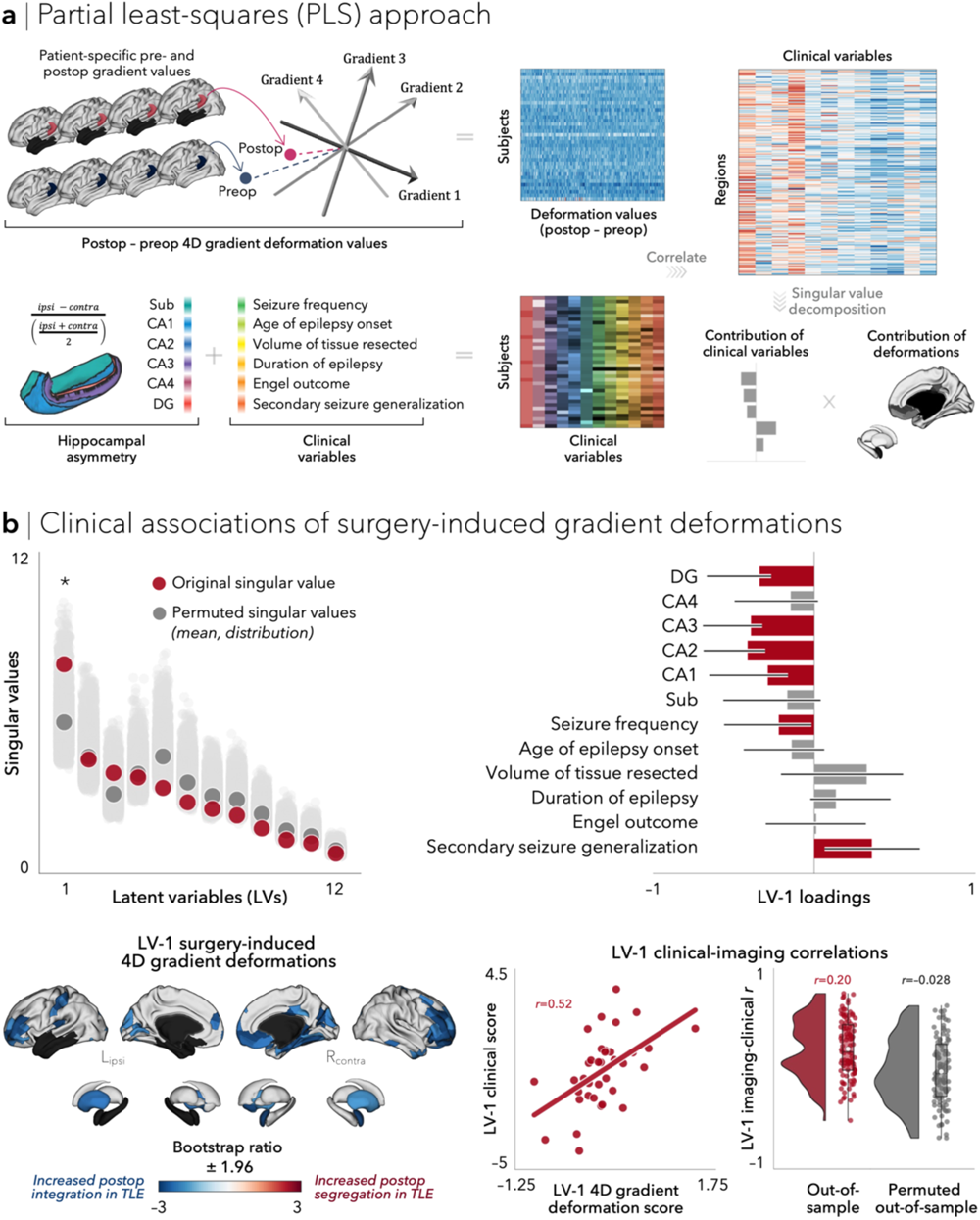
A clinical-imaging signature of surgery-induced connectome reorganization. (**a**) Partial least-squares (PLS) analysis was used to relate patient-specific pre-to postoperative 4D gradient deformation with clinical variables. These two sets of variables are correlated across patients and subjected to singular value decomposition, yielding multiple latent variables. Bootstrap resampling evaluated the contribution of individual variables to the latent variable, whereas permutation tests and cross-validation assessed the pairing of the 4D surgery-induced deformation patterns and clinical variables. (**b**) The first latent variable (LV-1) accounted for 44.65% of the covariance between the imaging and clinical data (*p* < 0.05; top left). The contribution of individual clinical measures is shown using correlations between patient-specific clinical scores and scores on the multivariate pattern (loadings; top right). Error bars indicate bootstrap-estimated SEs. The contribution of individual regions is shown using bootstrap ratios (interpretable as *z*-scores; bottom left). Patients who displayed increased postoperative connectome integration had greater ipsilateral hippocampal atrophy in CA1-3 and DG subfields, lower seizure frequency, and secondarily generalized seizures. This association was confirmed by projecting individual patient data onto the weighted patterns (*r* = 0.52; bottom middle). Correlations between 4D surgery-induced deformations and clinical scores in cross-validated held-out data (mean ± SD *r* = 0.20 ± 0.29) were higher than those from the permuted null model (mean ± SD *r* = –0.028 ± 0.33; bottom right).

## Discussion

Contemporary systems neuroscience is increasingly exploring advanced dimensionality reduction techniques to delineate the main organizational axes of the human connectome, and to derive potential clinical predictors.^48^ Here, we used connectome gradient mapping—an established dimensionality reduction method—to investigate local-global brain interactions in individuals with pharmaco-resistant TLE. We capitalized on this condition as a human lesion model to deepen our understanding of network-level consequences of focal lesions and their resection. We found that, before surgery, individuals with TLE showed an increased segregation of the ipsilateral temporal lobe (*i.e.*, the disease epicenter) from the rest of the brain. By tracking pre-to postoperative connectome alterations, we showed that resection of the ipsilateral temporal lobe led to brain reorganization involving areas near the resection site, but also marked re-integration of contralateral temporo-parietal regions with the rest of the brain. We further provided proof-of-concept evidence of an association between inter-individual variations in connectome reorganization following surgery and clinically salient features. Taken together, our results demonstrate the potential of gradient mapping to bridge different scales of human brain organization and allow the study of the brain’s structural plasticity in response to focal lesions and their eventual resection in young adults.

Core to our analytical framework was the estimation of connectome gradients that index the spatial arrangement of macroscale organization along continuous cortico-subcortical axes. This layout serves as a meaningful coordinate system to contextualize different MRI-derived measures with other markers of neural organization. For instance, such gradient approaches have been used to investigate the associations between macroscale brain organization and transcriptomic,^60^ synaptic,^61^ molecular,^62^ microstructural,^63,64^ and structure-function^9,65^ factors. Gradient mapping studies aiming to better understand brain changes observed in psychiatric and neurological conditions—including epilepsy—are also starting to emerge. Indeed, recent stepwise functional connectivity studies and applications of dimensional decompositions have described an imbalance of functional integration and segregation between sensory and transmodal systems in TLE.^66,67^ Our study applied gradient mapping for the first time to structural connectivity data in epilepsy. Using diffusion MRI, we demonstrated shifts in macroscale connectome organization implicating strategic regions such as the disease epicenter itself, as well as distant, yet direct, temporo-parietal connections, reflecting an increased segregation of these regions from the rest of the brain. Our findings further extend previous diffusion MRI studies that have focused on specific characteristics of white matter tracts either locally or globally,^25,68,69^ and instead simultaneously evaluated structural connectome changes both at the lesion site and in distant regions. Our approach provides evidence that a condition with a focal lesion to the mesiotemporal lobe presents with changes that extend well beyond the site of primary pathology, affecting directly and indirectly structurally connected macroscale networks. Debate persists over the precise mechanisms that underlie changes in these structural networks, but such changes may reflect excitotoxic effects from seizure activity or deafferentation of mesiotemporal connections.^70^ In this context, a possible explanation for the observed segregation of the ipsilateral anterior temporal cortex may relate to hyperconnectivity within the epileptogenic mesiotemporal network, together with hypoconnectivity of the epileptogenic mesiotemporal network to the rest of the brain, or a combination of both. Under this account, topological “isolation” of the ipsilateral temporal lobe may represent a core connectional change that may incur locally sustained hyperexcitability and increased susceptibility to seizure activity.^71^ On the other hand, hypoconnectivity between ipsilateral temporal lobe regions and all other regions may be considered as a compensatory mechanism through which epileptogenic activity is contained, abrogated, and prevented from spreading.^72,73^

A common surgical procedure to alleviate seizures in patients with medically-intractable TLE is anterior temporal lobectomy; *i.e.*, the resection of 4-4.5 cm of the temporal lobe (or up to 6.5 cm in case of non-dominant TLE).^21^ Here, we carried out longitudinal neuroimaging to track connectome reorganization before and after anterior temporal lobectomy. In our study, we showed that surgery-induced changes occur primarily in regions that were also affected preoperatively. The affected areas were bilateral, spanning regions near the resection site and contralateral temporo-parietal cortices. Albeit marginal, we showed evidence of postoperative connectome segregation of ipsilateral posterior temporal lobe regions, possibly pointing to ongoing Wallerian degeneration in nerve bundles disconnected after surgery. This process generally occurs in two phases, with acute dying back of axons within the first week postsurgery,^74^ followed by chronic myelin degradation lasting several months.^75^ With respect to the latter, there is ample evidence that ipsilateral temporal lobe fibers undergo irreversible Wallerian degeneration, with diffusion changes being apparent throughout the first year postsurgery.^75–77^ In line with these findings, a possible explanation for our observation of contralateral postoperative changes is the degeneration of commissural fibers connecting the two hippocampi and their afferent pathways.^23^ In contrast to preoperative alterations, changes in contralateral temporo-parietal cortices, which are expected to incur minimal downstream microstructural damage with surgery, were predominantly characterized by increased postoperative connectome integration. This shift in their connectivity profiles may be related to structural plasticity of the white matter in an active attempt to compensate for the resected grey matter and ongoing axonal/myelin degradation in homologous regions. Although such a relationship remains speculative, previous data have demonstrated that diffusion changes in contralateral fiber tracts fail to normalize following surgery.^76^ The absence of longitudinal data in controls in the current study may have hindered our ability to assess whether connectivity reorganization stabilizes, heals, or reorganizes over time relative to controls. Test-retest reliability studies of both diffusion MRI-derived connectomes^78^ and gradient-based frameworks^48^ have nevertheless shown high reliability in healthy adults.

Mining inter-individual variability in MRI data has been a cornerstone concept in neuroscience research.^79,80^ As a proof-of-concept, here we translated knowledge about connectome reorganization into individualized and clinically-meaningful taxonomies using latent clinical-imaging dimensions that link pre-to postsurgical shifts in connectome organization with heterogenous clinical measures. Our findings captured unique associations between connectome re-integration following surgery, primarily in bilateral fronto-occipital cortices, ipsilateral postcentral gyrus, and contralateral mesiotemporal regions, and greater ipsilateral hippocampal atrophy, lower seizure frequency, and presence of secondarily generalized seizures. As postsurgical seizure outcomes did not emerge as a significant contributor in our model, it is possible that a combination of both preoperative and postoperative features may better reflect seizure outcome, as suggested by a range of machine learning studies reporting associations between network measures and outcomes.^29,81–86^ Alternatively, outcome predictors may be identified from lower order latent variables, ideally with associated patterns of connectivity shifts centered on contralateral temporo-parietal and default-mode regions.^73^ Future studies assessing predictive features will ideally include multi-site data to assess predictive generalizability, enroll patients with longer-term seizure outcomes, and include both seizure and cognitive outcome measures to comprehensively assess prognostication of overall functioning and wellbeing. Nevertheless, the present results complement recent efforts to identify latent imaging-derived disease factors that not only variably co-express across TLE patients, but also have potential for individualized predictions.^87^

Taken together, these findings offer a new framework that can enrich our understanding of the effects of focal lesions and their eventual surgical resection on the macroscale organization of the human brain connectome. We dichotomized macroscale connectivity shifts before and after surgery in TLE, with the ipsilateral anterior temporal lobe—the target area for resection—being segregated from the rest of the brain prior to surgery. Secondary to surgery, contralateral temporo-parietal regions revealed connectivity shifts in a diametrically opposite direction, likely reflecting a compensatory response to restore or maintain structural connectivity. This data-driven connectome reorganization further accounted for a broad spectrum of clinical factors, potentially opening up new avenues for future neuroimaging studies to parse inter-individual differences in epilepsy and, consequently, may be translatable to individualized patient care. More broadly, a human lesion model, like the one described herein, offers a rare window into the building blocks of brain organization in both healthy and diseased states, and can ultimately allow causality to be inferred based on the effects of focal brain lesions in TLE.

## Data Availability

All data produced in the present study are available upon reasonable request to the authors

## Acknowledgments

S.L. acknowledges funding from Fonds de la Recherche du Québec–Santé (FRQ-S), the Canadian Institutes of Health Research (CIHR), and the Richard and Ann Sievers Neuroscience Award. B.Y.P. was supported by the National Research Foundation of Korea (NRF-2021R1F1A1052303; NRF-2022R1A5A7033499), Institute for Information and Communications Technology Planning and Evaluation (IITP) funded by the Korea Government (MSIT; No. 2022-0-00448, Deep Total Recall: Continual Learning for Human-Like Recall of Artificial Neural Networks; No. RS-2022-00155915, Artificial Intelligence Convergence Innovation Human Resources Development [Inha University]; No. 2021-0-02068, Artificial Intelligence Innovation Hub), and Institute for Basic Science (IBS-R015-D1). J.R. received support from the Canadian Open Neuroscience Platform (CONP) and CIHR. B.F. receives funding from FRQ-S (Chercheur-Boursier clinician Senior). A.N. acknowledges funding from FRQ-S. E.S. acknowledges funding from FRQ-S. G.S. acknowledges support from CIHR. B.M. acknowledges support from NSERC (Discovery Grant RGPIN 017-04265) and from the Canada Research Chairs Program (CRC). A.B. and N.B. were supported by FRQ-S and CIHR (MOP-57840, MOP-123520). Z.Z. was supported by the National Science Foundation of China (NSFC: 81422022; 863 project: 2014BAI04B05 and 2015AA020505) and the China Postdoctoral Science Foundation (2016M603064). B.C.B. acknowledges support from CIHR (FDN-154298, PJT-174995), SickKids Foundation (NI17-039), NSERC (Discovery-1304413), Azrieli Center for Autism Research of the Montreal Neurological Institute (ACAR), BrainCanada, FRQ-S, the Helmholtz International BigBrain Analytics and Learning Laboratory (Hiball), and the Canada Research Chairs program.

## Author contributions

S.L., Z.Z., and B.C.B contributed to the conception and design of the study; Y.W., Z.W, Z.Z, and R.L. contributed to the acquisition of data; S.L. and B.C.B contributed to data analysis and manuscript writing; revised and approved by all co-authors.

## Potential conflicts of interest

None to disclose.

## Data availability

Codes to conduct multimodal preprocessing (micapipe^40^), hippocampal segmentation (HippUnfold^56^), and statistical analysis (BrainStat^49^) are openly available.

## Notes

### Competing Interest Statement

The authors have declared no competing interest.

### Funding Statement

This study was funded by CIHR

### Author Declarations

Ethics committee/IRB of Jinling Hospital, Nanjing University School of Medicine gave ethical approval for this work.

