## Supplement 1 for "Connectome reorganization associated with temporal lobe pathology and its surgical resection"

### Supplementary Materials

| | **TLE** ($n$ = 37) | **HC** ($n$ = 31) |
|---|---|---|
| **Age** (mean ± SD) | 26.70 ± 7.62 | 27.29 ± 7.26 |
| **Sex** (male/female) | 18/19 | 14/17 |
| **Side of focus** (left/right) | 16/21 | – |
| **Age at onset** (mean ± SD) | 14.86 ± 7.77 | – |
| **Duration of illness** (mean ± SD) | 11.33 ± 7.37 | – |
| **Seizure frequency** | 8.21 ± 21.15 | – |
| **Secondarily generalized seizures** (presence/absence) | 31/6 | – |
| **Time between preoperative scan and surgery** (mean ± SD) | 5.09 ± 7.55 | – |
| **Time between surgery and postoperative scan** (mean ± SD) | 15.38 ± 15.92 | – |
| **Surgical outcome** (Engel I/Engel II-IV) | 26/11 | – |

**Table S1**. **Demographics of individuals with TLE and healthy controls**. Demographic breakdown of TLE and control cohorts, including age (in years) and sex. Clinical parameters in TLE, including side of seizure focus, age at onset of epilepsy (in years), duration of illness (in years), seizure frequency (seizures/month), and presence or absence of secondarily generalized seizures. Surgery-related information in TLE, including time between preoperative MRI scan and surgery (in months), time between surgery and postoperative MRI scan (in months), and postsurgical seizure outcome.

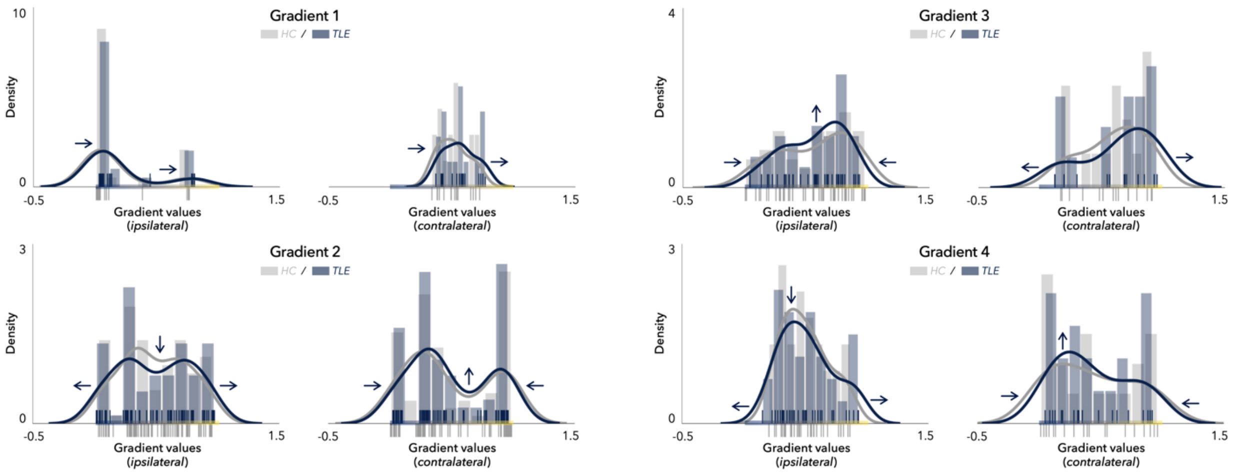


**Figure S1**. **Histograms of significant preoperative gradient changes**. Gradient-specific histograms (split into ipsilateral/contralateral) reveal three distinct patterns of alterations, namely: a shift (*e.g.*, Gradient 1), a contraction (*e.g.*, Gradient 2 contralateral), and an expansion (*e.g.*, Gradient 2 ipsilateral) of gradient values in TLE.

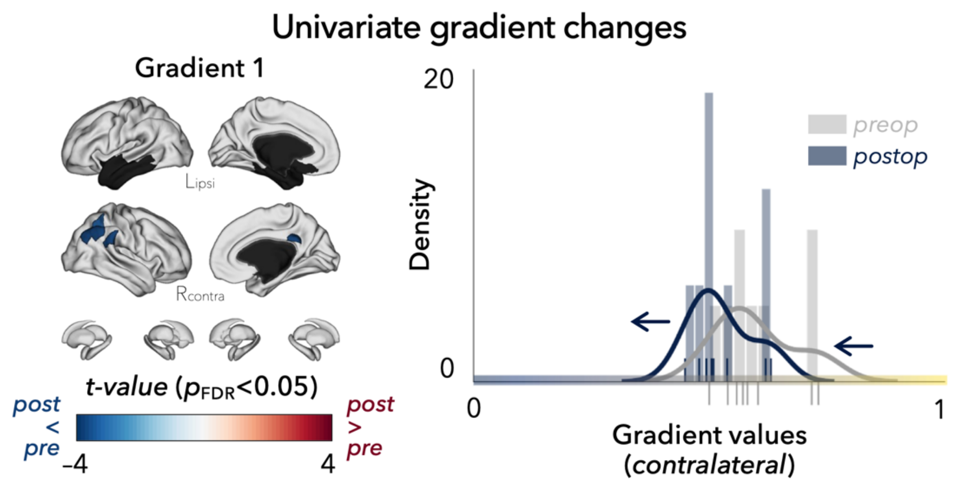


**Figure S2**. **Pre- to postoperative univariate gradient changes**. Univariate mixed effects models compared pre- to postoperative gradient values in individuals with TLE. Univariate linear models revealed significant pre- to postoperative changes in Gradient 1 only, affecting contralateral precuneus and parietal regions (all $p_{\mathrm{FDR}} < 0.05$). After surgery, gradient values in these regions were shifted towards the middle, more integrated positions along Gradient 1.

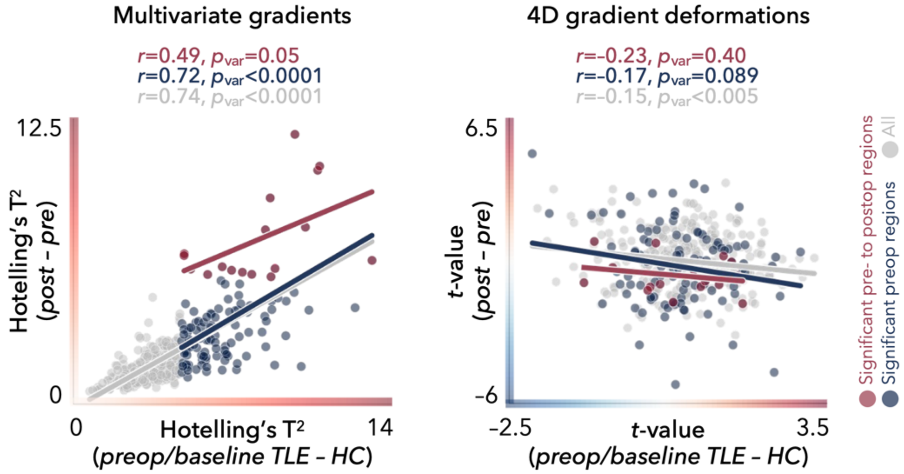


**Figure S3**. **Preoperative gradient changes constrain postoperative reorganization**. The degree of preoperative multivariate gradient changes (relative to controls) was spatially correlated with pre- to postoperative multivariate gradient changes across all regions ($r = 0.74$, $p_{\text{var}} < 0.0001$) and within areas of significant gradient changes (cross-sectional: $r = 0.72$, $p_{\text{var}} < 0.0001$, longitudinal: $r = 0.49$, $p_{\text{var}} = 0.05$). Moreover, negative spatial associations between preoperative and pre- to postoperative 4D gradient deformations were observed, albeit significant only when considering all regions ($r = -0.15$, $p_{\text{var}} < 0.005$).
